# Exploring the implemented guidelines for dyslipidemia treatment and care among nurses and physicians in Jordanian hospitals

**DOI:** 10.1101/2025.01.29.25321329

**Authors:** Rana K. bani Salameh, Ahmed M. Al-Smadi, Omar Gammoh, Abedalmajeed M. Shajrawi, Ala Ashour, Omar Alrfooh, Donna Fitzsimons, Taher Hatahet, Ammena Yahia Binsaleh, Sireen Abdul Rahim Shilbayeh

**Author notes:** Corresponding author (TH).

## Abstract

**Background:** Dyslipidemia is a major risk factor for ischemic heart disease in the world. The last updated ACC/AHA and ESC/EAS guidelines for dyslipidemia management aimed to control blood cholesterol and reduce cardiovascular risk. In Jordan, no national guidelines have been published to date, so these guidelines are commonly used. There are a few studies exploring the implementation of these guidelines in Jordan.

**purpose:** To explore the implementation of updated ACC/AHA and ESC/EAS guidelines for the treatment and care of dyslipidemia among nurses and physicians in Jordan.

**Method:** Semi-structured interviews (three focus group) with open-ended questions was conducted to gain deeper understanding of implemented practice and guidelines among nurses and physicians.

**Results:** Focus group interviews revealed that health care professionals try to implement specific guidelines and reach for optimal goals, but many barriers prevent them from reaching therapeutic goals, like workload, limited resources, and a lack of support from health institutions. They reported many suggestions that may enhance their ability to implement dyslipidemia management guidelines and wanted to improve specific guidelines in Jordan for dyslipidemia management.

**Conclusions:** Results indicated limited implementation of any dyslipidemia management guidelines among Jordanian nurses and physicians in their clinical practices.

## Introduction

Dyslipidemia is defined as “elevated Total Cholesterol (TC), low-density lipoprotein cholesterol (LDL-C), or triglycerides (TG); low high-density lipoprotein cholesterol (HDL-C); or a combination of these abnormalities.” ^1^ In 2019, WHO considered dyslipidemia a major risk factor for two causes of death in the world: ischemic heart disease (IHD) and stroke ^2^. In 2017, the WHO estimated the worldwide deaths from IHD and ischemic stroke at 3.9 million (3.7–4.2 million) for one type of dyslipidemia (high non-HDL cholesterol) ^3^. Each year, 4 million people in the United States and 1.5 million in the European Union die from cardiovascular disease (CVD), according to the European Society of Cardiology (ESC)^4^.

All deaths of CVDs accounted for 37% in Jordan in 2018^5^. In a Jordanian cohort research, the top risk factors for ischemic stroke were hypertension, diabetes, hyperlipidemia, and coronary artery disease ^6^. A national prevalence study on the lipid profile in Jordan in 2017 found that 44.3% had high TC, 41.9% had high TG, 75.9% had high LDL-C, and 59.5% had low HDL-C ^7^.

Physicians and nurses have a major role in treating and managing dyslipidemia ^8^.There are a few studies exploring the implementation of dyslipidemia management guidelines among nurses and physicians in Jordan. Halawani et al (2019) revealed the importance of physicians’ knowledge of the latest guidelines for increasing quality of patients care and would give more satisfactory results about lipids profile ^9^. Another study in Jordan reported a low level of knowledge about American College of Cardiology/American Heart Association (ACC/AHA) guidelines among Jordanian physicians^10^. Furthermore, another study conducted in Jordan showed the awareness of dyslipidemia (9.3%), the treatment (50.3%), and the control (25.4%) among adults in Jordan ^11^. The last updated ACC/AHA guidelines and European Society of Cardiology/ European Atherosclerosis Society (ESC/EAS) guidelines for dyslipidemia management aimed to control blood cholesterol and reduce cardiovascular risk ^12,13^. To the best of our knowledge, in Jordan, no national guidelines for the management of dyslipidemia have been published up to date, which presents a significant challenge for healthcare professionals. The absence of national guidelines leads to healthcare providers to rely on international guidelines, particularly those from the ACC/AHA and ESC/EAS guidelines which are commonly used. Due to the lack of prior research in this study area, this study was conducted to explore the implementation of updated (ACC/AHA) and (ESC/EAS) guidelines for the treatment and care of dyslipidemia among nurses and physicians in Jordan, to explore the level of treatment control for patients with dyslipidemia in Jordan and to explore the barriers and suggestions for implementing these guidelines among physicians and nurses in Jordan.

## Method

A qualitative study was conducted, contain three focus group semi-structured interviews. The study included two public Jordanian hospitals and one teaching hospital: King Abdullah University Hospital, Al Basheer Governmental Hospital, and Al Mafraq Governmental Hospital. The hospitals are located in two geographical areas in north and central Jordan.

### Sample

Three focus groups were conducted, consisting of at least one cardiologist, one internal medicine physician and four nurses for each group from public Jordanian hospitals. The sample included different types of health care professionals from four cardiac units (critical care unit, intensive care unit, Intermediate, and medical and surgical cardiac ward) within the cardiology and cardiac surgery departments in each of the three settings. Only included nurses and physicians with at least one year of experience in their units. The researcher conducted a meeting to invite individuals to participate in this study. The participants were selected using a purposive sampling method^14^.The small sample size in this study was determined by practical constraints, including time and resource limitations. Three hospitals were selected to ensure a diverse representation of healthcare settings. One central and one teaching hospital was included to provide in-depth data from a high-resource environment, reflecting large, specialized facilities. Another additional hospital from smaller, regional settings was chosen for comparison. Although the sample size is limited, it is sufficient to achieve the study’s exploratory goals and provide valuable insights into the impact of hospital resources on patient outcomes. Furthermore, this sample size allows for thorough data collection and sets the stage for larger studies in the future.

Additionally, The sample size, as defined by Creswell and Poth and Polit and Beck ^15^, is the number of study participants and is determined by the data saturation principles and the information needs of the research.

### Ethical consideration

Ethical approval was obtained from the Al al-Bayt University of Jordan and from the institutional review boards of each hospital that is included in this study (File 1). All participants were voluntary and anonymous. The consent form was written prior to data collection. Confidentiality was applied.

### Tools for data collection procedure

Focus group semi-structured interviews with physicians and nurses, the semi-structured interviews were conducted using an interview guide with open-ended questions derived from the literature (File 2). The questions focus on implemented practices and guidelines for dyslipidemia treatment and care among physicians and nurses in Jordan, as well as barriers and suggestions for implementing dyslipidemia guidelines. Focus groups were conducted in mixed groups with physicians and nurses, with one group from each hospital. Each group contained five to ten participants. A meeting was held with participants to form groups. All the participants were given information about focus groups and interview goals. Informed consent was obtained from all of them (File 3). All interviews were digitally recorded and transcribed verbatim. Each interview lasted approximately 20 minutes.

### Data collection procedure

The researcher contacted 20 healthcare professionals during their clinical work who meet inclusion criteria and asked them to participate in semi-structured, focus group interviews after fully explaining the current study. The consent forms were provided. The participants were asked for a suitable date and time to conduct the semi-structured interview after confirming their consent from. After obtaining demographic information, a semi-structured focus group tape-recorded interview was conducted in Arabic in a suitable quiet place within the hospital setting. The interviews started with the researcher presenting the study goals and the specific goal of the interview to the participants. The researcher then ensured a comfortable setting, set up the recording equipment, and started a general chat with participants, followed by an open-ended questions that were directed by the interview guide schedule. The average time for an interview was approximately 20 minutes, which was acceptable to the participants in the context of their clinical work and was suitable for reaching saturation. The first author conducted all interviews. Data were collected between 1th of June 2012 and 1th of August 2022.

### Thematic analysis

The main goal was to explore more details about implemented practices, barriers, and suggestions for guideline implementation among nurses and physicians. The data was analyzed using thematic analysis. The analysis was conducted by the researcher using the guidelines reported by Braun and Clarke’s approach^16^.Thematic analysis involves number of interrelated but distinct stages or steps which are (1) data immersion; (2) generating initial codes; (3) searching for themes;(4) reviewing themes and ;(5) defining and naming themes. The audio recording was a transcript; the researcher first translated the interviews into Arabic verbatim (word for word) and then translated them back into English. Then, the interviews’ data was organized, and each interview was given a number or code. Names and other personally identifying information were removed from the transcripts.

After organizing the data, the data immersion stage started by reading and rereading the whole interview transcripts, and re-listening to all or parts of the audio recording, the researcher became familiar with the whole interview. The preliminary coding stage was started by carefully reading the transcripts line by line, and a label or code was applied to anything that might be important. Then, the codes were compared, grouped together, and organized into categories around similar and interrelated ideas or concepts and at this stage the first initial coding was identified. The analysis then progressed to the systematic application of these codes, using the established categories and codes to index the entire data set. Categories were considered to be dense when no new open codes could be inferred from the data, indicating theoretical saturation^17^. Each code was assigned a number or abbreviation and directly written onto the transcripts. The next step involved generating themes by examining the coded data for patterns and commonalities. Themes were created by grouping similar codes that represented broader concepts or recurring ideas within the data. After that, in the reviewing stage, the developed themes were refined and compared with the original transcripts to ensure they were grounded in the actual data. Once the themes were refined and validated, the themes were completed for further definition and description of each theme’s content.

### Measures for ensuring trustworthiness

To enhance the rigor and quality of the findings, The researcher evaluated the trustworthiness of qualitative data^18^.

### Credibility

Credibility was enhanced by using member checking. The interview transcripts were given to the all participants, who were then invited to check the information for accuracy. They were asked to verify or elucidate any information in order to guarantee that their answers were appropriately recorded and depicted.

### Transferability

In order to address the transferability, the research was conducted in three different settings which represented the public health care sector in Jordan, with different health care providers from different cardiac units, which enhanced the generalizability of the findings.

### Dependability

Data were collected in the appropriate settings, with appropriate participants, and times to enhance the dependability of the study. To further ensure consistency, an audit trail was maintained, recording every stage of the investigation, including gathering data, analyzing it, and making decisions.

### Conformability

The researcher explicitly described the study methods and procedures in order to help the reader to follow the actual consequence of how data were collected, processed, analyzed, and interpreted. The entire research process, as well as each step of the data analysis and findings were discussed and reviewed with the study team and experts (supervisors and a qualified nurse).

## Results

As stated in the methodology part focus group interviews were tape recorded after a meeting with participants to create groups and after receiving consent forms. An interview guide with open-ended questions was used (File 3). Table 1 show the main three themes emerging from interview data. In this section, each theme is given separately with supporting quotations.

**Table 1.**
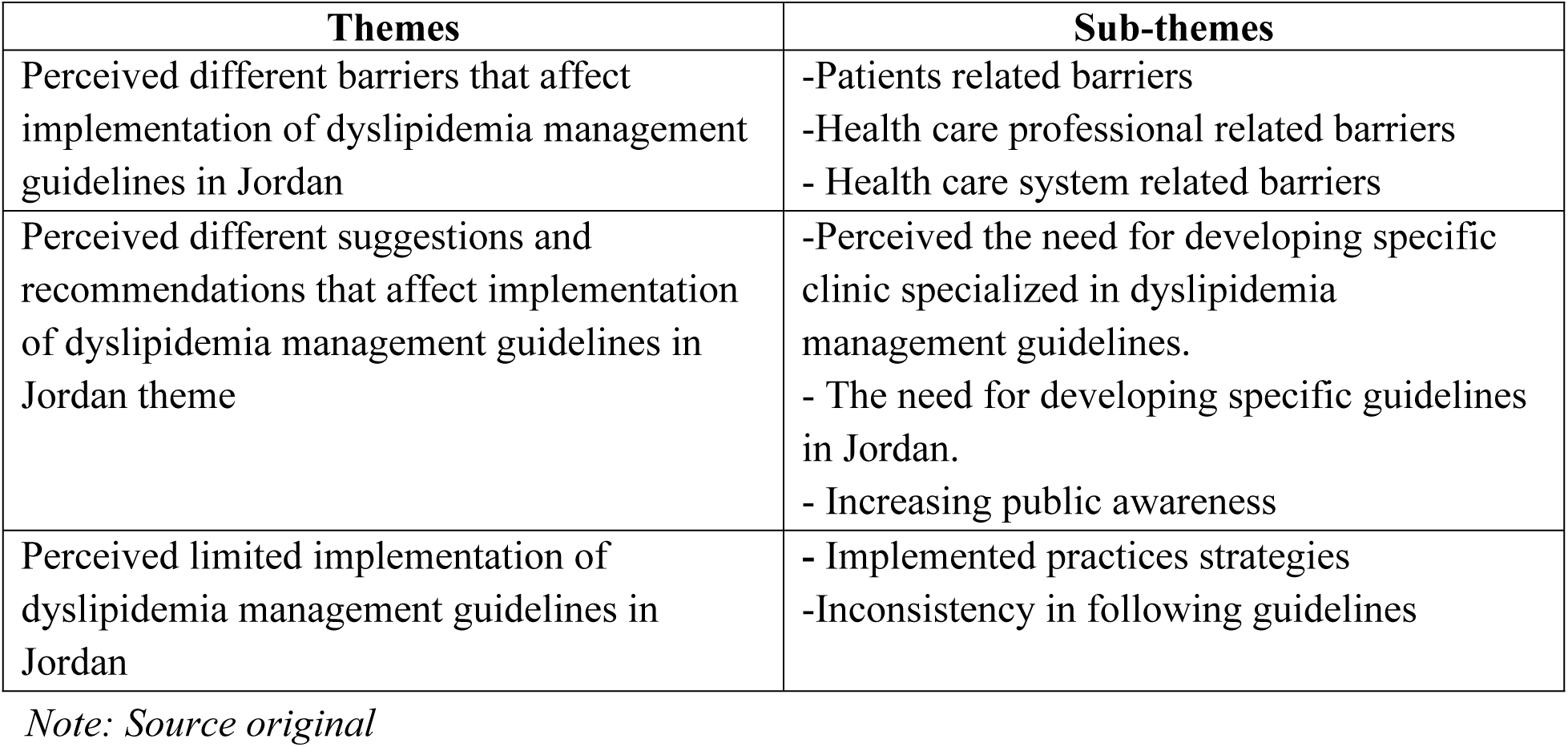
Themes and sub-themes emerging from the interview data.

### 1) Perceived different barriers that affect implementation of dyslipidemia management guidelines in Jordan theme

The theme included the following sub-themes: patients related barriers; health care professional barriers; and health care system related barriers.

#### A: Patients related barriers: sub-theme

The majority of participants suggested that patient noncompliance, no follow-up, psychological status, financial status, and cultural issues were the most common patient-related barriers that affect the implementation of dyslipidemia treatment and care guidelines in Jordan.

Participants reported that patients had a limited compliance with their prescribed medications and instructions from their doctors, and there was no follow-up to check their lipid profiles. Patients follow an unhealthy lifestyle.

> “*One of the most barriers that affect dyslipidemia management guidelines implementation in Jordan is the lack of compliance, because the patients think that medication is just for one month. They do not know that they have to continue their medication forever. They do not make lipid profile test, do not eat healthy food or even follow diet and healthy lifestyle*.” Cardiologist, number 9.

The majority of participants, both physicians and nurses, stated that patients did not follow up even after we advised them and that changing their lifestyle was difficult. Patients don’t like to wait long hours for their appointment and prefer to buy their medication from a private pharmacy without follow-up.

> “*Even if I give advice to my patients, large number of them do not follow it; particularly, if it is related to change their lifestyle*”. Registered nurse, number 11.

#### B: Health Care Professional Related Barriers: Sub-Theme

Many participants reported that the most significant health care professional-related barriers affecting the implementation of dyslipidemia treatment and care guidelines in Jordan are a lack of knowledge, poor communication, and a lack of follow-up.

“*We have Lack of knowledge about dyslipidemia management guidelines. There is no training course for nurses and physicians”*. Registered nurse, number 3.

“*There is no good communication between patients with physicians and nurses* .*”* Internal medicine, number 1.

#### C: Health Care System Related Barriers: Sub-Theme

Many participants stated that the most health-care system-related barriers to implementing dyslipidemia management guidelines in Jordan are a lack of interest and a lack of a follow-up system, limited resources, a heavy workload, and a lack of support for health institutions.

> “*The main barrier that face us is the shortage of the Number of physicians and nurses*”. Internal medicine, number 1.

> “*Some of medication not available or they have high cost, other medication sometimes available and sometimes not. That will prevent us to implement guidelines*”. Registered nurse, number 4

The majority of participants stated that work overload and time pressure were two of the most important barriers among physicians and nurses that prevent them from implementing dyslipidemia management guidelines in their hospital.

> “*We don’t have enough time to implement dyslipidemia management guidelines in our hospitals and patient culture .*” Registered nurse, 3.

### 2) Perceived different suggestions and recommendations that affect implementation of dyslipidemia management guidelines in Jordan theme

The majority of participants suggested the need for developing specific clinic specialized in dyslipidemia management guidelines, need for developing specific guidelines in Jordan and increase public awareness by making free lipid profile, education program, enhance follow up system

> “*One of the solutions is to make clinical center to treat hyperlipidemia in Jordan like KHCC. When five hundred patients come to my clinic and some of them have hyperlipidemia, it is very difficult to offer injections. So, these patients should be transferred to dyslipidemia center. In Holland, they transfer the patients who have LDL above specific limit*”. Cardiologist, number 9.

### 3) Perceived limited implementation of Dyslipidemia management guidelines in Jordan theme

Majority of participants reported they don’t apply any guidelines in their hospitals, but they appeared to be familiar with the dyslipidemia management guidelines.

> “*We don’t follow any guidelines for dyslipidemia management in our hospital*”. Registered nurse, number 2

> “*We have a guideline, but we don’t apply it because we don’t have correct follow up*”. Cardiologist, number 9

All of participants reported they apply their experience to manage patients with dyslipidemia.

> “*I start to manage patient’s life style, if not response I give anti lipid medication for patients with stroke, DM, HTN. I don’t follow specific; I just follow my experience*”. Internal medicine, number 1.

## Discussion

The qualitative findings revealed that the majority of participants did not use any recommendations in their clinical practices, while some attempted to follow easier and more up-to-date rules. However, all participants stated that they manage patients with dyslipidemia based on their experience, and some nurses stated that the guidelines constitute medical management rather than nursing management. Previous findings suggested that the dyslipidemia care guidelines were not implemented and followed by all participants, and they relied on their clinical experience to handle patients with dyslipidemia.

The current study’s findings revealed a low level of adherence to the implementation of dyslipidemia management guidelines in Jordanian physicians’ and nurses’ clinical practices. These findings are consistent with recent research, which found that physicians and clinical pharmacists in Saudi Arabia and Jordan had a low level of understanding of the 2013 ACC/AHA guidelines recommendations 10 19 for dyslipidemia care . However, both studies found that physicians had a poor degree of knowledge of this rule, implying that the guidelines were not followed in their clinical practices.

Many challenges to applying clinical guidelines have been identified in previous research^20^. Our findings revealed numerous challenges that could explain why there is such a low level of adoption among Jordanian physicians and nurses. The most common barriers to adoption of dyslipidemia management principles were a lack of resources, work overload, time constraints, a lack of support, a lack of understanding, and lack of knowledge. Furthermore, our findings are consistent with earlier research, which found numerous impediments to implementing any guidelines in a literature review and a comprehensive mixed method studies review ^21, 22,23^.

The results of this study align with several previous findings that suggest patient noncompliance is a significant challenge in managing chronic diseases like dyslipidemia. Many participants noted that patients often fail to follow prescribed treatments, and there is a lack of follow-up to monitor lipid profiles. This is consistent with research by Taniguchi et al. (2024), who found that inadequate follow-up care and poor medication adherence were among the primary factors influencing treatment effectiveness^24^.

Additionally, the study found that patients tend to follow unhealthy lifestyles, which may exacerbate their conditions and hinder guideline adherence. This observation echoes the findings of a randomized study, which found that lifestyle modifications, including diet and exercise, are often neglected by patients despite being key components of treatment for dyslipidemia^25^. Furthermore, psychological status is a crucial barrier to adherence to treatment regimens, particularly among patients experiencing mental health challenges such as depression or anxiety. This highlights the importance of integrating mental health care into the management of chronic diseases, as integrated behavioral healthcare (IBH) has been shown to improve treatment adherence by addressing both physical and mental health needs rates^26^. In addition, Our results emphasize financial resources as a major patient-related barrier. This finding is consistent with the fact that many people are unable to access healthcare due to insufficient insurance, financial constraints, inability to pay, and the absence of clinics or medical providers in their area^27^.

In addition to patient-related factors, health care professionals also play a key role in the successful implementation of guidelines. Some participants noted that lack of proper training among health professionals could contribute to suboptimal care. This aligns with findings from Hamaideh (2017), who observed that insufficient training and limited knowledge among healthcare providers often lead to inconsistent application of treatment guidelines^28^. Furthermore, the lack of specialized training in dyslipidemia management was identified as a critical gap in the healthcare system, where clinicians are not consistently trained to recognize and implement the guideline effectively^29^. In addition, Youssef et al (2018) noted that healthcare professionals in many countries lack up-to-date knowledge about current clinical guidelines, which directly affects the quality of care provided^30^. Moreover, poor communication between healthcare providers and patients contributes to a lack of patient engagement, which, as shown by Tiwary et al (2019), leads to poor treatment adherence and suboptimal health outcomes. Inadequate patient-provider communication can create significant barriers to effective chronic disease management, especially in conditions like dyslipidemia, where long-term adherence to lifestyle changes and medications is required^31^.

The study also highlighted significant healthcare system-related barriers, such as lack of interest, limited resources, heavy workload, and lack of support for health institutions. These systemic challenges play a crucial role in hindering the effective implementation of dyslipidemia management guidelines in Jordan. Such barriers are consistent with broader studies on healthcare systems in developing countries, where similar issues are frequently encountered. For example, one study found that healthcare systems in low-resource settings, including Jordan, often struggle with insufficient infrastructure and lack of institutional support, both of which are essential for the successful implementation of clinical guidelines. Without the necessary structural and financial backing, it becomes difficult for healthcare providers to consistently apply treatment protocols, leading to suboptimal care^28^.

The lack of a structured follow-up system is another critical issue that exacerbates the situation, and this has also been discussed in several previous studies^32^. For instance, a retrospective cohort study showed that merely 18% of people diagnosed with dyslipidemia during the health checkup sought follow-up care from a physician within a 6-month period, this ultimately prevents the healthcare system from achieving desired health outcomes for dyslipidemia management^33^. Our findings emphasized that the lack of organized follow-up mechanisms, coupled with the scarcity of resources such as diagnostic tools, medications, and trained personnel, results in inconsistent application of treatment guidelines^34^. Thus, regular follow-up is essential for monitoring patients’ progress, adjusting treatment plans, and ensuring adherence to prescribed medications.

Moreover, heavy workload and the lack of resources for healthcare professionals were consistently cited as major barriers by participants in this study. This finding reflects the experiences documented by a scoping review, which noted that healthcare providers in overloaded systems frequently do not have sufficient time, energy, or support to follow clinical guidelines rigorously. In busy healthcare environments, professionals are often forced to prioritize urgent care over preventive or long-term management, which leads to the neglect of guidelines that are crucial for chronic disease management.^35^ Furthermore, The workload was consistently emphasized as a significant obstacle that hindered physicians from adhering to the clinical practice guidelines^36^.

Furthermore, the lack of support for health institutions from the government was identified as a critical factor. Several studies highlighted how insufficient funding, inadequate facilities, and limited governmental backing result in health institutions being unable to maintain the necessary resources to implement guidelines effectively^34,35,36^. Without a supportive policy environment, healthcare facilities struggle to implement the guidelines, thus undermining the healthcare system’s ability to meet public health goal^35,37^.

Based on the findings, several key suggestions were identified to improve the implementation of dyslipidemia management guidelines in Jordan. Advocating for additional resources and financial support to promote equitable healthcare access is comparable to a study conducted in Boston, Massachusetts, which emphasized the need for financial assistance^38^. Establishing specialized clinics and training for dyslipidemia management would provide focused care, control for cholesterol and improve patient adherence. One study concluded that cholesterol control in IHD patients was influenced by the type of training activity undertaken by physicians during the implementation of the clinical guidelines, which led to achieving a good level of control, in line with the recommendations of the guidelines^39^. Additionally, our participants suggest that national guidelines tailored to the local context are necessary to address cultural and resource-related factors. This is consistent with a scoping review in the literature, which emphasized factors related to the guidelines, such as simplifying the guidelines, providing easy access, and focusing on the design and development of the guidelines^35^. In addition, our participants reported that offering free lipid profile tests could enhance early detection and patient engagement. Additionally, public health campaigns aimed at raising awareness of dyslipidemia management guidelines align with existing literature on the promotion and dissemination of clinical practice guidelines, as well as on the awareness of CPGs and confidence in their importance^40,41^. Our study suggests that implementing a structured follow-up system is crucial to ensure ongoing care and adherence, enhance regular monitoring, improve patient outcomes, and facilitate guideline implementation. These findings align with studies that emphasize the importance of consistent leadership and the commitment of team members^42^. Finally, continuous training for healthcare providers is crucial to ensure they remain current with the latest guidelines, aligning with the facilitators identified in the systematic review^40^.

One of the most concerning findings of this study is the perceived limited implementation of dyslipidemia management guidelines across healthcare institutions in Jordan. While participants expressed awareness of the guidelines, the majority reported that they were not actively using them. This reflects a broader issue in the region where, despite the existence of evidence-based guidelines, the adoption of these guidelines is often inconsistent. Our findings are similar to those highlighted in other studies regarding the limited implementation of guidelines, with a list of barriers that can influence this process^34^. Thus, the disconnect between knowledge of guidelines and their implementation can be attributed to the barriers discussed above, including inadequate training, poor communication, and systemic challenges such as limited resources and follow-up mechanisms.

There are several strengths in the current study. It is one of the first studies conducted in Jordan to explore how physicians and nurses implement the updated American College of Cardiology/American Heart Association (ACC/AHA) and European guidelines for dyslipidemia treatment. This research provides valuable insights into how international guidelines are applied in the local context. The findings will be crucial for developing specific, localized dyslipidemia management guidelines in Jordan. By identifying key barriers and facilitators, the study offers a foundation for improving clinical practice and patient outcomes.

While this study provides valuable insights findings in the implementation of dyslipidemia management guidelines, there are a number of limitations that must be discussed in this study. Firstly, our sample size is representative, but if we were involved more, it would be even more representative. Secondly, our settings were not involved in all the healthcare sectors; they only involved two governmental hospitals, and one educational hospital. Thirdly, BMI, blood pressure, and blood test data were gathered from patients’ medical records in three locations with varying standards rather than using standardized measurements. Fourthly, our study didn’t include pharmacist, patients and dietitian. Finally, our tools are newly developed, further research are needed to evaluate psychometrics properties.

The findings of this study have significant implications for both policy and practice. To improve the implementation of dyslipidemia management guidelines, healthcare policymakers in Jordan should prioritize training healthcare professionals, particularly in the areas of updated guidelines and effective patient communication. Additionally, healthcare institutions should be equipped with the resources needed to implement the guidelines effectively. Investment in specialized clinics, public awareness campaigns, and the establishment of a national follow-up system are also critical to overcoming the existing barriers. Moreover, the development of Jordan-specific guidelines, which account for local cultural and economic factors, would facilitate a more tailored and effective approach to dyslipidemia management. By addressing these barriers and implementing the suggested recommendations, there is a significant opportunity to improve the quality of care for dyslipidemia patients in Jordan and other similar regions.

## Conclusion

The results of the current study showed a low level of implementation of any dyslipidemia management guidelines among Jordanian nurses and physicians in their clinical practices. The nurses and physicians were responded difficulty they face in implementation Besides, they gave suggestions that may help them implement dyslipidemia management guidelines. Hence, future research should focus on the development and evaluation of context-specific guidelines, the impact of specialized clinics, the use of digital tools for follow-up, and the strengthening of healthcare infrastructure.

Investigating cultural, socioeconomic, and behavioral factors that influence patient adherence, as well as examining effective public health campaigns, will also be critical in advancing dyslipidemia management in Jordan and other similar settings.

## Data availability

The data that support the findings of this study are available from the Corresponding author upon reasonable request.

## Data Availability

All relevant data are within the manuscript and its Supporting Information files.

## Acknowledgment

The abstract of this study was presented in association of cardiovascular nursing and allied professions conference in the following link (https://doi.org/10.1093/eurjcn/zvad064.071)

## Supporting information

**S1. File. Ethical approval.**

**S2. File. Open ended questions for focus group.** Focus group semi-structured interview questions for physicians and nurses.

**S3. File. Focus group consent form.**

**S1.Table 1. Themes and sub-themes emerging from the interview data.** The main three themes emerging from interview data.

## References

1. Schwinghammer TL, Dipiro JT, Ellingrod VL, et al. Pharmacotherapy Handbook, 11e. McGraw. 2021.

2. 2019. URL https://www.who.int/news-room/fact-sheets/detail/thetop10causes-ofdeath.

3. Taddei C, Zhou B, Bixby H et al. Repositioning of the Global Epicentre of Non-Optimal Cholesterol. Nature 2020; 582: 73– 77.

4. Statistics2017 C and Ehnheart, 2022. URL https://ehnheart.org/cvd-statistics/cvd-statistics-2017.html.

5. 2018. URLhttps://www.who.int/nmh/countries/jor_en.pdf.

6. Alawneh KZ, Qawasmeh A, Raffee M et al. A snapshot of Ischemic Stroke Risk Factors, Sub-Types, and Its Epidemiology: Cohort Study. Ann Med Surg 2020; (59): 101– 105.

7. Abujbara M, Batieha A, Khader Y et al. The Prevalence of Dyslipidemia Among Jordanians. J Lipids 2018; .

8. Ding R, Ye P, Zhao S et al., 2017.

9. Halawani AFM, Alahmari ZS, Asiri DA et al. Diagnosis and Management of Dyslipidemia. Study Journal of Global Health 2019; 10(4).

10. Rababa’h AM, Mustafa R, Rabab’ah MA et al. Evaluation of Physician’s Knowledge in Jordan about the ACC/AHA Blood Cholesterol Guidelines. International Journal of Clinical Practice 2021; 75(3).

11. Pengpid S and Peltzer K. Prevalence, Awareness, Treatment, and Control of Dyslipidemia and Associated Factors Among Adults in Jordan: Results of A National Cross-Sectional Survey in 2019. Preventive Medicine Reports 2022; 28.

12. Grundy SM, Stone NJ, Bailey AL et al. PCNA Guideline on The Management of Blood Cholesterol: A report of the American College of Cardiology/American Heart Association Task Force on Clinical Practice Guidelines. Journal of the American College of Cardiology 2019; 73(24): 285–350.

13. Mach F, Baigent C, Catapano AL et al. 2019 ESC/EAS Guidelines for the Management of Dyslipidemias: Lipid Modification to Reduce Cardiovascular Risk. Atherosclerosis 2019; 290: 140–205.

14. Barton A. Research Methods: A Practical Guide for the Social Sciences. British Journal of Criminology [Internet]. Oxford University Press; 2012 Sep 1;52(5):1017–1021. Available from: https://academic.oup.com/bjc/article-abstract/52/5/1017/470134

15. Matthews R, Ross E. Research Methods: A practical guide for the social sciences. Pearson Education Ltd, 2010.

16. Clarke V, Braun V. Thematic analysis. J Posit Psychol. 2017;12(3):297–298. doi:10.1080/17439760.2016.1262613.

17. Charmaz K. Grounded theory as an emergent method. In: Hesse-Biber SN, Leavy P, editors. Handbook of emergent methods. New York: Guilford Press; 2008. p. 155–172.

18. Shenton AK. Strategies for ensuring trustworthiness in qualitative research projects. Educ Inf. 2004;22:63–75.

19. Zaitoun MF, Iflaifel MH, Almulhim LA et al. Awareness of Physicians and Clinical Pharmacists About ACC/AHA Guidelines for Dyslipidemia Management: A Cross-SectionalStudy. Journal of Pharmacy & Bioallied Sciences 2019; 11(2): 181–186.

20. Haesler E and Carville K. Implementing the 2019 International Guideline: consultation on barriers, facilitators, challenges and resource needs. Wound Practice& Research: Journal of the Australian Wound Management Association 2020; 28(2): 90– 96.

21. Hall AM, Scurrey SR, Pike AE et al. Physician-Reported Barriers to Using Evidence-Based Recommendations for Low Back Pain in Clinical Practice: A Systematic Review and Synthesis of Qualitative Studies Using the Theoretical Domains Framework. Implementation Science 2019; 14(1): 1– 19.

22. Mathieson A, Grande G and Luker K, 2019. URL 10.1017/S1463423618000488.

23. Correa VC, Lugo-Agudelo LH, Aguirre-Acevedo DC, et al. Individual, Health System, and Contextual Barriers and Facilitators for The Implementation of Clinical Practice Guidelines: A Systematic Metareview. Health Research Policy and Systems 2020; 18(1): 1–11.

24. Taniguchi Y, Iwagami M, Sugiyama T, Kuroda N, Yamaoka T, Inokuchi R, Suzuki A, Watanabe T, Irie F, Tamiya N. Factors associated with non-attendance at a follow-up visit for dyslipidemia identified at health checkups: A retrospective cohort study in a Japanese prefecture. JMA J 2024;7(4):518–528.

25. Pamplona-Cunha H, Rosini N, Caetano R, Machado MJ, da Silva EL. Lifestyle Intervention in Reducing Cardiometabolic Risk Factors in Students with Dyslipidemia and Abdominal Obesity: A Randomized Study. International Journal of Cardiovascular Sciences [Internet]. Int J Cardiovasc Sci; 2021 Jun 30; Available from: https://ijcscardiol.org/article/lifestyle-intervention-in-reducing-cardiometabolic-risk-factors-in-students-with-dyslipidemia-and-abdominal-obesity-a-randomized-study

26. Buchanan G, Berge JM, Piehler TF. Integrated behavioral health implementation and chronic disease management inequities: an exploratory study of statewide data. BMC Primary Care. BioMed Central 2024 Aug 14;25(1).

27. 1. Duncombe DC. A multi-institutional study of the perceived barriers and facilitators to implementing evidence-based practice. Journal of Clinical Nursing 2018 Jan 15;27(5– 6):1216–26. doi:10.1111/jocn.14168

28. Hamaideh SH. Sources of Knowledge and Barriers of Implementing Evidence-Based Practice Among Mental Health Nurses in Saudi Arabia. Perspectives in Psychiatric Care [Internet]. Perspect Psychiatr Care; 2017 Jul 1;53(3):190–198. Available from: https://pubmed.ncbi.nlm.nih.gov/28681446/

29. Duncombe DC. A multi-institutional study of the perceived barriers and facilitators to implementing evidence-based practice. J Clin Nurs 2018 Mar;27(5-6):1216–1226. doi: 10.1111/jocn.14168. Epub 2018 Jan 15. PMID: 29149462.

30. Youssef N, Alshraifeen A, Alnuaimi K, Upton P. Egyptian and Jordanian nurse educators’ perception of barriers preventing the implementation of evidence-based practice: A cross-sectional study. Nurse Education Today [Internet]. Churchill Livingstone; 2018 May 1;64:33–41. Available from :https://www.sciencedirect.com/science/article/pii/S0260691718300637

31. Tiwary A, Rimal A, Paudyal B, Sigdel KR, Basnyat B. Poor communication by health care professionals may lead to life-threatening complications: examples from two case reports. Wellcome Open Res 2019 Jan 22;4:7. doi: 10.12688/wellcomeopenres.15042.1. PMID: 31448336; PMCID: PMC6694717.

32. Menon S, Smith MW, Sittig DF, Petersen NJ, Hysong SJ, Espadas D, et al. How context affects electronic health record-based test result follow-up: A mixed-methods evaluation. BMJ Open 2014 Nov;4(11). doi:10.1136/bmjopen-2014-005985

33. Taniguchi Y, Iwagami M, Sugiyama T, Kuroda N, Yamaoka T, Inokuchi R, Suzuki A, Watanabe T, Irie F, Tamiya N. Factors associated with non-attendance at a follow-up visit for dyslipidemia identified at health checkups: a retrospective cohort study in a Japanese prefecture. JMA J 2024;7(4):518–528.

34. Abboud J, Rahman AA, Shaikh N, et al. Physicians’ perceptions and preferences for implementing venous thromboembolism (VTE) clinical practice guidelines: a qualitative study using the Theoretical Domains Framework (TDF). Arch Public Health 2022;80:52. doi:10.1186/s13690-022-00820-7.

35. Fischer F, Lange K, Klose K, Greiner W, Kraemer A. Barriers and Strategies in Guideline Implementation-A Scoping Review. Healthcare (Basel) 2016 Jun 29;4(3):36. doi: 10.3390/healthcare4030036. PMID: 27417624; PMCID: PMC5041037.

36. Gittus M, Chong J, Sutton A, Ong ACM, Fotheringham J. Barriers and facilitators to the implementation of guidelines in rare diseases: a systematic review. Orphanet J Rare Dis 2023 Jun 7;18(1):140. doi: 10.1186/s13023-023-02667-9. PMID: 37286999; PMCID: PMC10246545.

37. De Hert S, Paula–Garcia WN. Implementation of guidelines in clinical practice; barriers and strategies. Current Opinion in Anaesthesiology. 2024 Jan 17;37(2):155–62. doi:10.1097/aco.0000000000001344

38. Barnes SJ, Na Y, Drainoni M-L, Linas BA, Bosch NA, Tamlyn AL. Barriers and facilitators to conducting human subjects research at a safety net institution from the perspective of researchers. PLOS ONE 2025 Jan 8;20(1). doi:10.1371/journal.pone.0313530

39. Forcadell Drago E, Dalmau Llorca MR, Aguilar Martín C, Ferreira-González I, Hernández Rojas Z, Gonçalves AQ, López-Pablo C. Impact of Implementing a Dyslipidemia Management Guideline on Cholesterol Control for Secondary Prevention of Ischemic Heart Disease in Primary Care. Int J Environ Res Public Health 2020 Nov 19;17(22):8590. doi: 10.3390/ijerph17228590. PMID: 33228008; PMCID: PMC7699273.

40. Zhou P, Chen L, Wu Z, Wang E, Yan Y, Guan X, Zhai S, Yang K. The barriers and facilitators for the implementation of clinical practice guidelines in healthcare: an umbrella review of qualitative and quantitative literature. J Clin Epidemiol 2023 Oct;162:169–181. doi: 10.1016/j.jclinepi.2023.08.017. Epub 2023 Aug 30. PMID: 37657616

41. Bierbaum M, Rapport F, Arnolda G, Nic Giolla Easpaig B, Lamprell K, Hutchinson K, Delaney GP, Liauw W, Kefford R, Olver I, Braithwaite J. Clinicians’ attitudes and perceived barriers and facilitators to cancer treatment clinical practice guideline adherence: a systematic review of qualitative and quantitative literature. Implement Sci 2020 May 27;15(1):39. doi: 10.1186/s13012-020-00991-3. PMID: 32460797; PMCID: PMC7251711.

42. Correa VC, Lugo-Agudelo LH, Aguirre-Acevedo DC, Contreras JAP, Borrero AMP, Patiño-Lugo DF, Valencia DAC. Individual, health system, and contextual barriers and facilitators for the implementation of clinical practice guidelines: a systematic metareview. Health Res Policy Syst 2020 Jun 29;18(1):74. doi: 10.1186/s12961-020-00588-8. PMID: 32600417; PMCID: PMC7322919.

43. R Bani Salameh, A Al-Smadi, N Eshah, A Musa, O Al-Rawajfah, M Jarrah, Exploring the implemented guidelines for dyslipidemia treatment and care among nurses and physicians in Jordanian hospitals, *European Journal of Cardiovascular Nursing*, Volume 22, Issue Supplement_1, August 2023, zvad064.071, 10.1093/eurjcn/zvad064.071

